# Modeling COVID19 mortality in the US: Community context and mobility matter

**DOI:** 10.1101/2020.06.18.20134122

**Authors:** Sarah F. McGough, Ryan W. Gan, Robert Tibshirani, Anne-Marie Meyer

**Affiliations:** Genentech, Inc., South San Francisco, CA, USA; Department of Statistics, Stanford University, Stanford, CA, USA; Department of Biomedical Data Sciences, Stanford University, Stanford, CA, USA; F. Hoffmann-La Roche, Ltd., Basel, Switzerland; Department of Epidemiology, University of North Carolina at Chapel Hill, Chapel Hill, NC, USA

## Abstract

The United States has become an epicenter for the coronavirus disease 2019 (COVID-19) pandemic. However, communities have been unequally affected and evidence is growing that social determinants of health may be exacerbating the pandemic. Furthermore, the impact and timing of social distancing at the community level have yet to be fully explored. We investigated the relative associations between COVID-19 mortality and social distancing, sociodemographic makeup, economic vulnerabilities, and comorbidities in 24 counties surrounding 7 major metropolitan areas in the US using a flexible and robust time series modeling approach. We found that counties with poorer health and less wealth were associated with higher daily mortality rates compared to counties with fewer economic vulnerabilities and fewer pre-existing health conditions. Declines in mobility were associated with up to 15% lower mortality rates relative to pre-social distancing levels of mobility, but effects were lagged between 25-30 days. While we cannot estimate causal impact, this study provides insight into the association of social distancing on community mortality while accounting for key community factors. For full transparency and reproducibility, we provide all data and code used in this study.

**One-sentence summary:** County-level disparities in COVID19 mortality highlight inequalities in socioeconomic and community factors and delayed effects of social distancing.

## Main Text

The United States (US) has become an epicenter for the coronavirus disease 2019 (COVID-19) pandemic, with over 2 million confirmed cases and 115,000 confirmed deaths as of June 15, 2020. However, impact of the disease across the US has been unequal, with documented geographic heterogeneities in cases and deaths and excess burden in certain subgroups of the population. Clinical reports have noted the increased incidence and severity of COVID-19 among individuals with diabetes, obesity, and chronic lung conditions (*1, 2*). Other influential community factors include social and economic vulnerabilities (*3, 4*), referred to in the epidemiologic literature as “social determinants of health” (*5*). Many of these social determinants of health have been shown to be closely correlated with race/ethnicity and specifically racial inequalities (*6*). Recent data from Chicago, New York, and Louisiana indicate that COVID-19 is disproportionately afflicting minority communities, primarily African American and Hispanic groups; in Louisiana, for instance, African Americans make up 32% of the population yet account for over 50% of deaths to date (*7*). However, population-level clinical and social determinants of health are only one component of the COVID-19 pandemic.

For example, social distancing policies have been adopted by state and local governments at different points during the outbreak, and have also been shown to impact COVID-19 growth rates (*8*). However, there is evidence to suggest that some communities initiated social distancing before formal implementation of policies (*9*), and may have relaxed prior to the official releases (*10*). Aggregated mobility data can capture these changes over time and offer a richer resolution for the continuum of community-level behavioral modifications (*11*). With economic and political pressures to release restrictions, it is critically important to assess how community-level sociodemographics and mobility play a role in COVID-19 mortality in order to develop effective response strategies.

Our study combines both community context and social distancing effects and evaluate COVID-19 mortality rates over time across US counties. By simultaneously incorporating population-level comorbidities, sociodemographic makeup, and social distancing over time, our approach identifies vulnerable communities at higher risk of mortality. Our study also quantifies the independent associations between declining mobility and subsequent reductions in mortality rates as well as mortality from community-level factors.

We investigated 24 counties surrounding 7 major metropolitan areas in the US that exhibit heterogeneities in sociodemographic characteristics and comorbidities (Figure 1A) as well as differences in timing of the adoption of social distancing over the course of the outbreak (Figure 1B). The metropolitan areas - Chicago, Detroit, Los Angeles, New Orleans, New York City, San Francisco, and Seattle - are similar to those examined in the CDC MMWR on COVID-19 and mobility (*12*). They were selected for their substantial size in COVID-19 cases and deaths, heterogeneity in social distancing adoption, and length of outbreak available to study. Counties were studied up to and including May 13, 2020, and the epidemiologic profile of each county is detailed in Table S1. We observed strong correlations between county-level characteristics including comorbidities and demographics (Figure S1, Table S2). To address these correlations, we applied principal components (PC) analysis to collapse potential COVID-19 county-level risk factors into four main representative constructs that explain 80% of the variation in sociodemographics across counties: “health and wealth,” “housing inequality” “old age and clinical care,” and average daily “PM_2.5_” (Figures S2, S3; described in Supplementary Materials, Materials and Methods). Together with data on daily human mobility, the principal components generated in this data pre-processing step were then used as predictors in a model for COVID-19 mortality.

**Figure 1.**
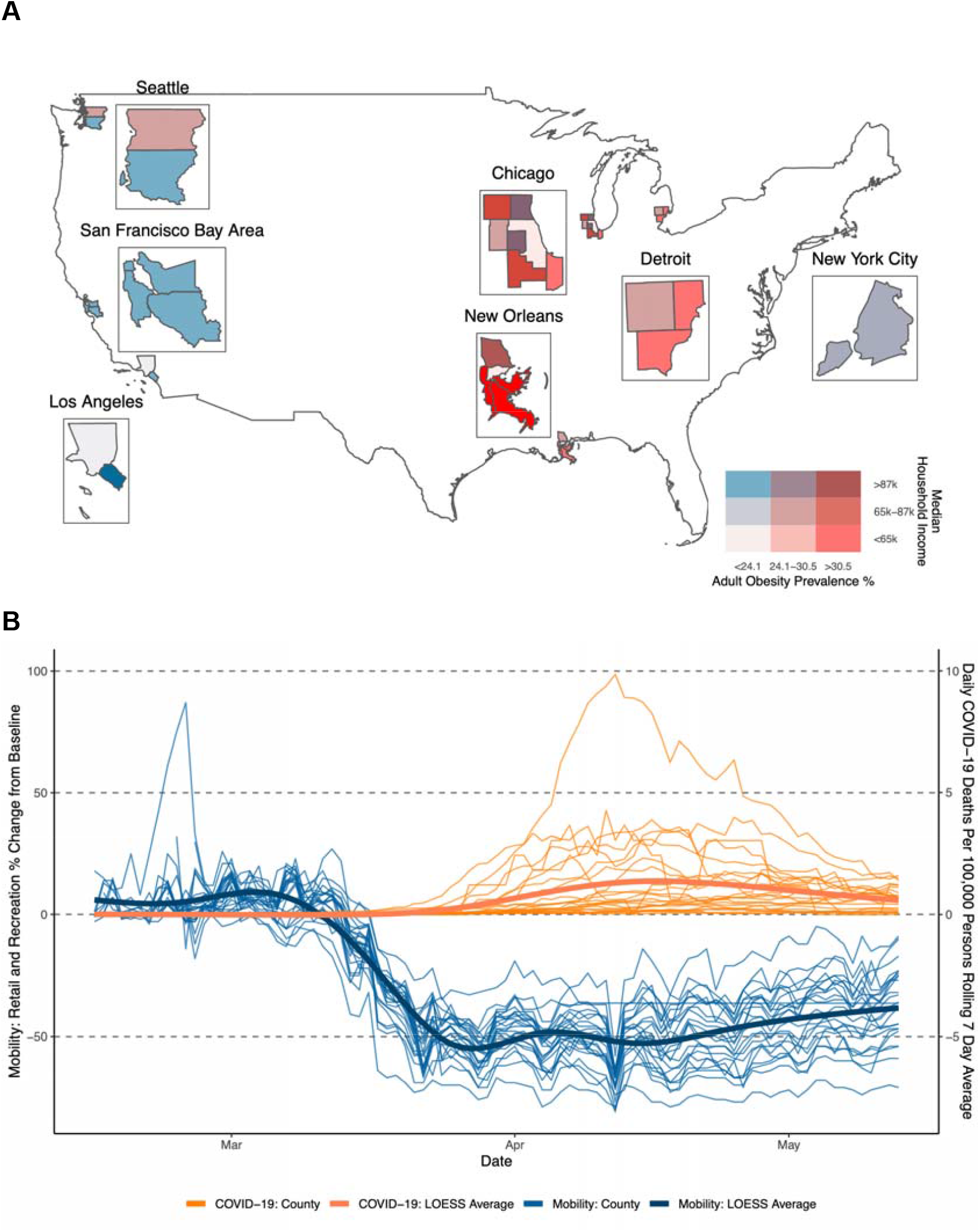
(A) Median household income (USD) and obesity prevalence (%) of the 24 study counties in 7 major metropolitan areas. (B) Google retail and recreational mobility (% change from Jan 3-Feb 6) (blue lines) and daily COVID-19 deaths per person (orange lines) across 24 counties over February 15, 2020 to May 13, 2020. In (B), study area counties are represented by finer lines and the broader lines represent the LOESS average across all counties over time. Daily COVID-19 deaths per person are represented as the 7-day rolling average, and baseline mobility is defined as the period between January 3 and February 6, 2020.

We used a generalized additive model (GAM), a flexible modeling approach (*13*), which uniquely and simultaneously addresses three key challenges not fully addressed in current literature: First, it allows for the quantification of a nonlinear relationship in COVID-19 mortality rates over time, while accounting for county-specific temporal evolution of COVID-19 deaths in these 24 counties. Second, it allows for quantification of the effect of the four PC county-level sociodemographic constructs. Third, it allows for quantification of the lagged relationship of social distancing over time using a penalized distributed lag framework, which can account for the time delay that occurs between infection and mortality (*14*). Here, we discuss the two major elements of the GAM model: 1) county-level sociodemographics and 2) social distancing as measured by changes in county-level mobility.

Economic vulnerabilities and poor baseline health indicators (“health and wealth”) had the strongest associations with COVID-19 mortality rates across counties, contributing nearly 10 times as much explanatory power to the model compared to the next PC predictor, “housing inequality” (F-statistic= 31.7 for “health and wealth” compared to 3.3 for “housing inequality”). Higher prevalence of comorbidities was collectively associated with higher COVID-19 daily mortality rates, including diabetes, obesity, and adult smoking (Figure 2; Figure S3). In contrast, higher median household income, higher levels of college education, and lower unemployment rates were collectively associated with lower COVID-19 daily mortality rates (Figure 2; Figure S3). These effect estimates also incorporate any changes in social mobility over time.

**Figure 2.**
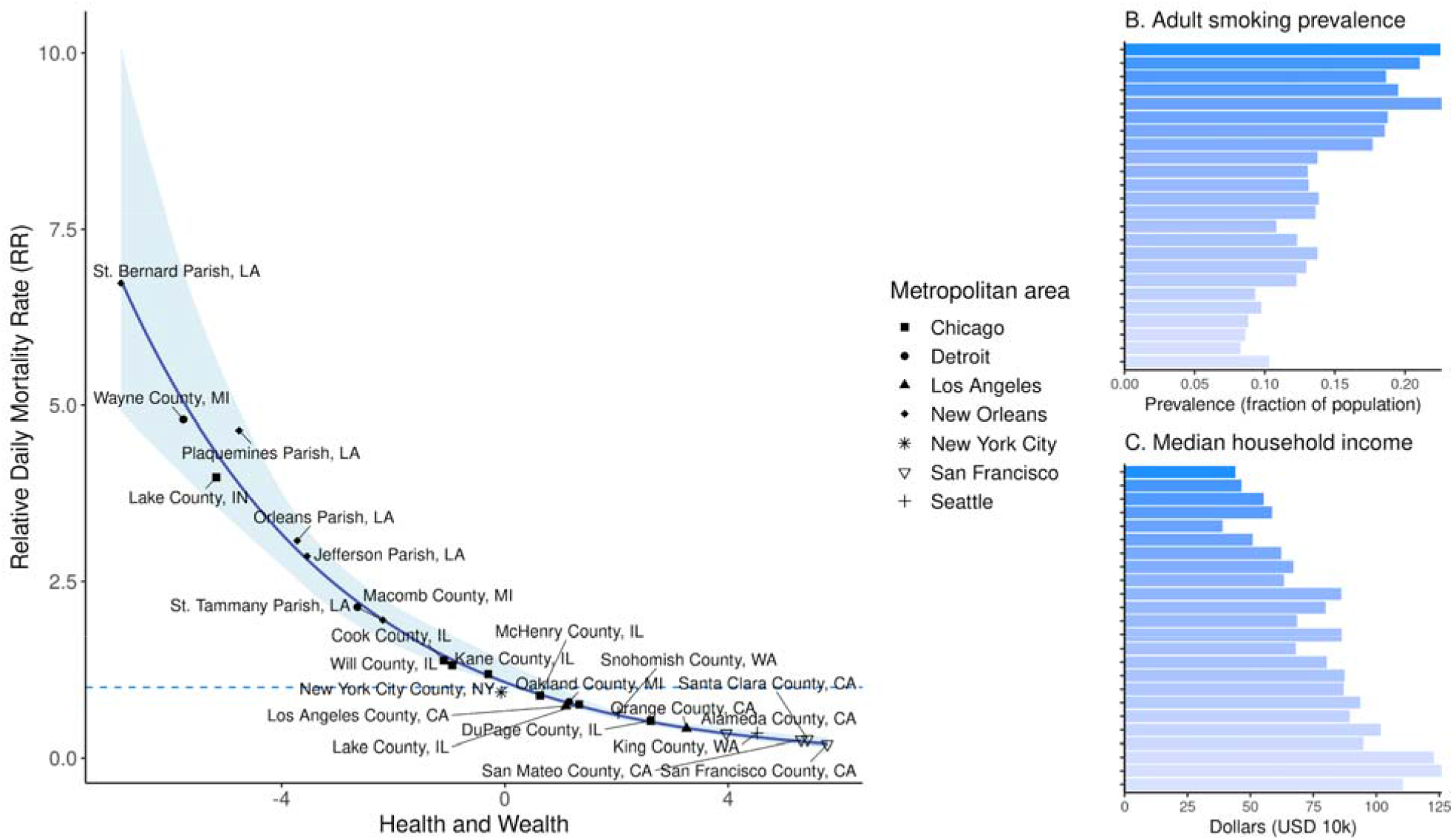
(A) Average estimated effects of “health and wealth” on daily mortality rate. For each county, we demonstrate with bar charts two sociodemographic predictors contained in the “health and wealth” principal component: (B) adult smoking prevalence and (C) median household income (in USD). 95% bias-corrected percentile intervals (shaded region) around the model estimate (blue line) are shown for 10,000 parametric bootstrap replicates in (A). From top to bottom, counties in (B) and (C) are ordered from lowest-to-highest “health and wealth” score.

Mortality rates associated with “health and wealth” were between 5 to 35 times higher in New Orleans-surrounding counties (St. Bernard, Jefferson, St. Tammany, and Plaquemines Parishes) compared to California Bay Area counties (San Francisco, San Mateo, Santa Clara, and Alameda Counties) after accounting for mobility. Though, St. Bernard, a smaller parish near New Orleans, experienced highly fluctuating numbers of deaths, resulting in wider uncertainty at the tails of mortality rate estimates (Figure 2).

It is important to note race/ethnicity was not implicitly included as an independent variable in county-level sociodemographic constructs. Rather, we included measures such as racial segregation, proportion non-native English speakers, income, and housing quality measures (Table S2), which represent the underlying community influences that contribute to vulnerable racial and ethnic minority groups. Communities that scored high on variables contributing to racial and ethnic subgroup vulnerabilities were at exceptionally high risk of COVID-19 mortality in this analysis, such as Detroit and New Orleans-area counties (Figure 2; Figure S3). Our results are complementary to current case and clinical reports that document a disproportionate amount of severe cases and deaths in disadvantaged populations, primarily African American and Hispanic communities (*4, 15*–*17*) and those populations with chronic underlying health conditions (*18*) including diabetes, hypertension, and obesity.

There has been considerable debate on the second component of our model, namely, the impact and effectiveness of social distancing on the COVID-19 outbreak. We used Google’s human mobility data (*19*) as a proxy for social distancing, as it objectively reflects the localized changes in movement early on in the outbreak before state and local social distancing policies were put into place. Because social distancing policies were issued in response to exponential growth in the epidemic, we observed a paradoxical relationship between the declining trajectory of mobility against a rising trajectory of COVID-19 deaths (Figure 1B), also observed in other populations (*20*). In this analysis, we controlled for the time evolution of the outbreak and modeled mobility as multiple distributed lags of 0 to 30 days to account for this real-time trend. This framework allows for a sufficient number of lag days without over-parameterizing the model.

Our model shows that reduced mobility in all study counties was associated with steady, linear declines in mortality rates over a 30-day period, after controlling for county-level principal components. Moreover, we found that it may take approximately 25 to 30 days to observe reductions in mortality rates following declines in mobility from baseline (reference point: 0% change in mobility, Figure 3). In the beginning of the observed epidemic, declines in mobility occur concurrently with a rise in mortality rates (Figure 1B), which is why studying the delayed (lagged) effect of mobility is critical to capture associations over time. The largest reductions in mortality rates were observed at the highest levels of social distancing (i.e. the highest reductions in mobility). For example, mortality rate reductions of 13% (95% CI: 7%, 17%) and 15% (95% CI: 9%, 24%) after 30 days were associated with decreases in mobility of 50% and 80%, respectively, compared to more modest mortality rate reductions after 30 days associated with decreases in mobility of 25% and 10% (Figure 3). Conversely, increasing mobility by 10% was associated with a small, but steady increase in mortality rate over the 30-day lag period.

**Figure 3.**
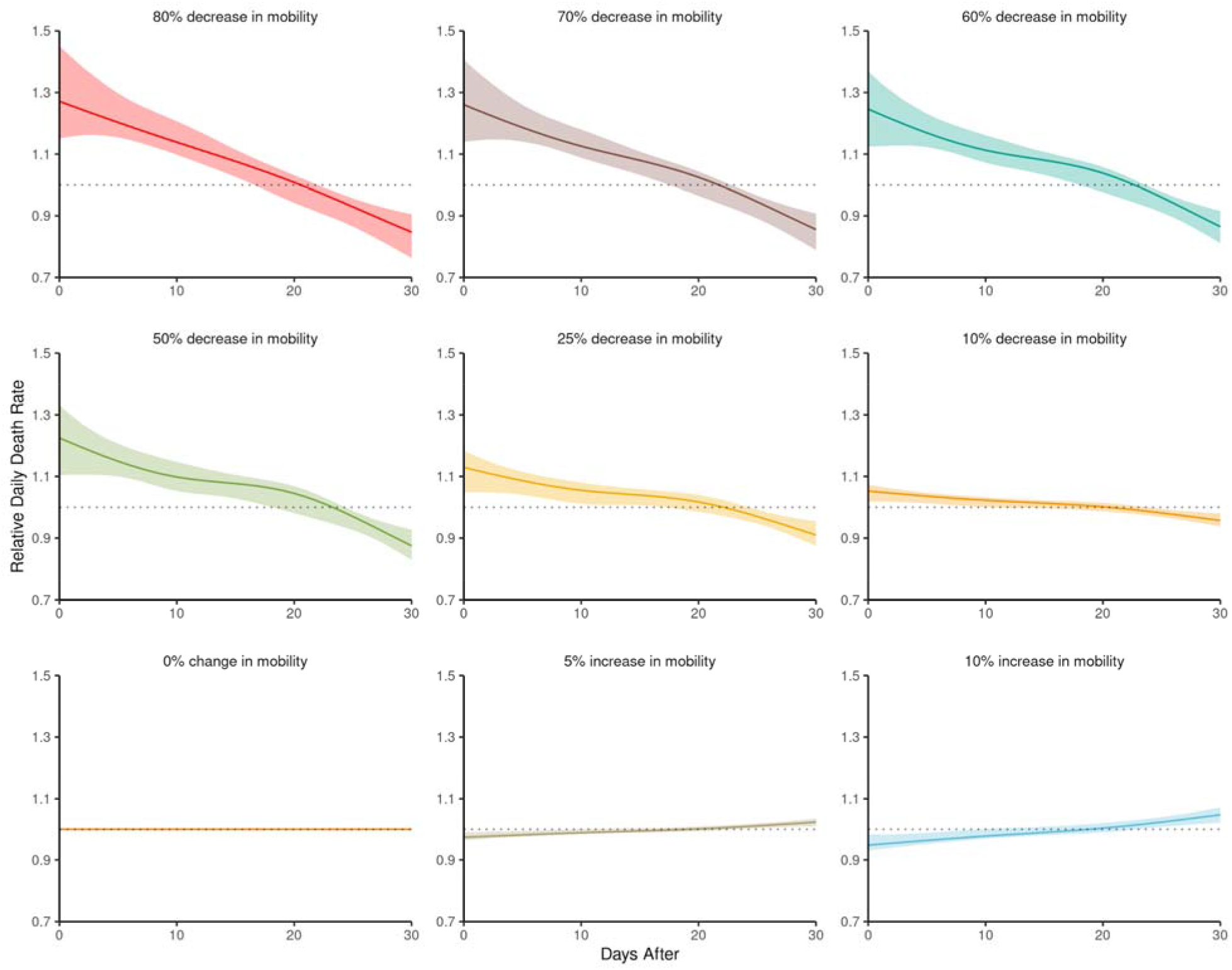
Associations between human mobility (% change from Jan 3-Feb 6) and daily mortality rates, at multiple lags (0-30 days after change in mobility), estimated across 24 study counties using penalized distributed lags. Mobility data were obtained from Google and represent the percent change from baseline (Jan 3-Feb 6) in retail and recreational mobility. 95% percentile intervals (shaded region) around the model estimate (thick line) are shown for 10,000 parametric bootstrap replicates.

It is possible that these relationships will change or strengthen over time as the outbreak persists, providing longer time periods in which to observe changes in mobility and lagged effects. However, these findings suggest that the relationships between mobility and mortality rates are complex, and it can take at least four weeks to observe notable reductions in deaths. These findings are consistent with our hypothesis that as communities in the US return to pre-social distancing levels of mobility, increases in mortality rates will likely be observed around four weeks later. Most importantly, these findings suggest that mortality rates in the initial days following social policy implementations may appear paradoxically higher compared to rates pre-social distancing, and take time to decline.

Sensitivity analyses were conducted to assess the robustness of findings around the principal components and lagged mobility. Results were similar when we: 1) conducted our statistical analysis using two different lagged modeling approaches for mobility, 2) excluded New York City because of its unusually high disease burden, and 3) truncated individual counties to the same period of observation time (Figures S4-S8).

Our results demonstrate it is critical to study COVID-19 in a broader context. By combining community-level variables and mobility data, we are able to understand their relative impact at a population level. As more data on COVID-19 emerge, evidence continues to mount on the sociodemographic disparities of the COVID-19 outbreak in the US and globally (*4, 15, 21*). As observed by Chung and colleagues (*16*), the scientific community is witnessing a “socioeconomic gradient” in the COVID-19 outbreak, in which disadvantaged communities are at higher vulnerability to COVID-19 infection and death. These social determinants of health are well described in the broader health literature (*22*), but have not been well elucidated at a population-level in the COVID-19 pandemic. The recent study by Chin and colleagues (*23*) outlines multiple characteristics that can be used to assess county-level vulnerability to COVID-19 to help plan and respond. Our analysis contributes a complementary contextual and statistical component, highlighting the intertwined risk factors of income, education, and pre-existing health issues and quantifying their associations with high mortality rates in vulnerable counties while accounting for declines in mobility.

This analysis also adds to the growing body of evidence on the relationship between mobility and COVID-19 (*20, 24, 25*), but takes a step further to contextualize the evolution of the outbreak at the county-level in the US. We show that social distancing was associated with reductions in COVID-19 mortality rates of up to 15% after 25-30 days, relative to baseline (Figure 3). Conversely, at current observed trajectories, reverting to pre-social distancing mobility was associated with an increase in mortality rates around 25-30 days later. Considering a maximum incubation period of roughly 14 days (*26*), followed by a period of illness or hospitalization prior to death, these estimates align with current understanding of COVID-19 transmission and progression.

Our analysis relies on population-level measures to draw these associations, and is not intended for drawing causal inferences or conclusions. It is important to acknowledge the limitation of population-level analyses, which could be biased by confounding from unmeasured variables and should not be extrapolated to individuals or be interpreted as individual risk.

Because policy and interventions are often targeted to a community level, we sought to explore factors potentially influencing COVID-19 mortality at a county-level, including social distancing, underlying health conditions, health access, income and income inequality, racial segregation, air pollution, and other sociodemographic factors. Modeling these factors collectively, their explanatory value and variance could be used to provide insight to local and state governments for policy evaluation and response planning.

This flexible, time series modeling framework can be extended to investigate other time-varying and time-invariant relationships of interest with COVID-19 mortality. The distributed lag approach enables time-continuous confounding adjustment (e.g. adjusting for the multiple lagged effects of mobility). Additional lagged terms at the same temporal and spatial resolution may be added to the existing model to explore other potential risk factors including meteorological variables and air pollution. And, as more data related to the pandemic become available, it will be possible to account for additional mitigating factors unavailable at this time, including other public health actions (e.g. wearing masks, contact tracing) and indicators of health system preparedness (e.g. hospital surge capacity, ventilators). Finally, the current analysis focuses on counties surrounding major metropolitan areas, and may not capture risk factors relevant to rural communities in the US. To better understand the generalizability of these results nationally and even globally, an extension of this work to additional locations is an important next step.

To the best of our knowledge, this analysis is the first to differentiate communities at-risk for COVID-19 mortality and, in the context of these factors, assess the relationship between social distancing and reductions in mortality over time. We identified potential community-level risk factors of COVID-19 mortality, including pre-existing health conditions (smoking, diabetes, obesity) and economic vulnerability (income, unemployment), and demonstrated the delayed effect of social distancing. Altogether, this analysis was made possible by the availability of open-access data from our sources including *The New York Times*, Google, University of Wisconsin Population Health Institute, Centers for Disease Control and Prevention, and Centers for Medicare and Medicaid Services. To promote “open science” and transparency in research, especially pertaining to the COVID-19 pandemic, we present a fully reproducible code repository with links to data and all scripts to replicate and extend this analysis at: https://github.com/phcanalytics/covid19_epi_model.

## Data Availability

All data and code used for this analysis can be found or downloaded at https://github.com/phcanalytics/covid19_epi_model.

## Acknowledgements

We thank the Roche COVID-19 task force for prioritizing this research, and the leadership at Roche and Genentech for supporting this scientific effort for the benefit of society. We also thank the broader “TPP2” teams within Diagnostic Information Solutions and Personalized Health Care for their collaboration and support on data, engineering, and meaningful scientific discussions on how to best study the COVID-19 pandemic and develop critical insight. We also thank N. Pal for code review and G. Simpson for his input on our use of the “mgcv” R package.

## Funding

R.T. is a paid consultant of Roche.

## Author contributions

S.F.M, R.W.G., R.T., and A.M. designed the study. S.F.M. and R.W.G. collected and processed data. S.F.M. and R.W.G. executed the statistical analyses. S.F.M. wrote the original draft of the manuscript. S.F.M, R.W.G., R.T., and A.M. edited and reviewed the manuscript. All authors read and approved the manuscript.

## Competing interests

The authors declare no competing interests.

## Listing of Supplementary Materials

**Materials and Methods**

**References (27 - 32)**

**Table S1 - S2**

**Fig S1 - S10**

## Notes

### Competing Interest Statement

The authors have declared no competing interest.

